# A cross-sectional study of the mismatch between telecommuting preference and frequency associated with psychological distress among Japanese workers in the COVID-19 pandemic

**DOI:** 10.1101/2021.05.20.21257516

**Authors:** Souhei Otsuka, Tomohiro Ishimaru, Masako Nagata, Seiichiro Tateishi, Hisashi Eguchi, Mayumi Tsuji, Akira Ogami, Shinya Matsuda, Yoshihisa Fujino, for the CORoNaWork Project

## Abstract

**Objective:** To examine how the mismatch between telecommuting preference and telecommuting frequency was associated with psychological distress during the COVID-19 pandemic.

**Methods:** Data from 33,302 workers throughout Japan were obtained using an Internet survey. Among 33,302 participants, 20,395 who telecommuted were included in the analysis. Participants’ telecommuting preference and frequency during the COVID-19 pandemic were determined using a questionnaire. Psychological distress was assessed using Kessler 6 (K6).

**Results:** Among participants who did and did not prefer to telecommute, those who telecommuted four or more days per week had an OR of psychological distress of 0.67 (p<0.001) and 1.87 (p=0.001), respectively, compared with those who rarely telecommuted.

**Conclusions:** The association between telecommuting and psychological distress differs depending on telecommuting preference.

## Introduction

Telecommuting has rapidly gained traction in many countries around the world including Japan during the coronavirus disease 2019 (COVID-19) pandemic. Following the World Health Organization (WHO)’s declaration of a pandemic on March 11, 2020, COVID-19 continued its rapid spread via the movement of people from urban areas in Japan, leading the government to declare its first state of emergency on April 16, 2020.^1^ In addition to urging companies to limit unnecessary trips out of the office and to avoid the “Three Cs” (closed spaces with poor ventilation, crowded places with many people nearby, close-contact settings such as close-range conversation), the government and local authorities also advised companies to promote telecommuting.^2^ This policy, which forced companies to take measures to balance infection control and business continuity, led most companies to rapidly adopt telecommuting. Before the COVID-19 pandemic, an estimated 20% of companies had telework systems in place and less than 10% of individuals were telecommuting. By the time the state of emergency had been extended nationwide, the proportion of companies adopting telecommuting had risen to about 60%.^3^

Although the impact of telecommuting on health remains unclear, telecommuting is suspected to have both positive and negative effects on health. In terms of positive effects, telecommuting reduces the burden of daily commuting and adds more flexibility to schedules. Telecommuting also improves work-life balance and job satisfaction.^4^ Workers who choose to telecommute experience fewer adverse health effects such as illness and disability.^5^ Telework environments are reportedly quieter, associated with reduced work interruptions and improved privacy, as well as better concentration, all of which contribute to workers’ health.^6^ However, negative health effects of telecommuting include psychological effects such as loneliness, isolation, and depressed mood^7–9^ and effects on the musculoskeletal system.^10–12^ With the spread of remote work and instructions to refrain from going out, there is a concern that individuals will feel disconnected to others as their involvement in the local community declines, leading to increased loneliness and anxiety.^13^ In addition, anxiety related to work management and communication, and difficulty separating one’s work life from one’s private life are sources of mental burden.^14^ Further, telecommuting is linked to concerns about eye strain and aggravation of stiff shoulders and back pain due to poor posture and sitting for long periods of time.^15^

Mental health has surfaced as a major public health issue during the COVID-19 pandemic. More than one-third (36.6%) of residents who experienced lockdown have reportedly experienced mild to moderate psychological distress.^16^ While the impact of telecommuting on psychological burden is not fully understood, we hypothesize that telecommuting is a potential risk factor for mental health. In particular, a mismatch between telecommuting preference and telecommuting frequency may impart a mental burden on telecommuters because telecommuting, when it was initiated under the state of emergency declaration, was adopted as a result of a forced public request rather than being based on workers’ wants. This is in contrast to cases in the past, when telecommuting was typically conducted based on workers’ wish to do so. While the Japanese government promoted telecommuting as a way to improve life-work balance before the COVID-19 pandemic, only a few companies had introduced it. Now, however, telecommuting, which has been rapidly adopted in response to the COVID-19 pandemic, is being implemented by companies in many industries and regions, and of all sizes.^17^ Therefore, the mismatch between telecommuting preference and telecommuting frequency is a new challenge for occupational health.

However, it is not known how a mismatch between telecommuting preference and telecommuting frequency affects psychological distress. We hypothesized that individuals who prefer to telecommute during the COVID-19 pandemic would feel less psychological distress, while those who prefer not to telecommute would feel more psychological distress with an increase in the frequency of telecommuting. This study examined how the mismatch between telecommuting preference and telecommuting frequency was associated with psychological distress during the COVID-19 pandemic.

## Methods

### Study Design and Participants

We performed a cross-sectional Internet monitor study from December 22 to 26, 2020, during Japan’s third wave of COVID-19 infection. The study protocol is reported in detail elsewhere.^18^ Briefly, we obtained data from workers who were employed at the time of the survey. Participating workers were selected based on region, job type, and sex. Since COVID-19 infection rates vary widely from region to region, we adopted regional sampling in this study.

Among 33,302 participants in the survey, 6,266 were excluded for providing fraudulent responses, defined as a markedly short response time (≤6 minutes), markedly low body weight (<30 kg), markedly short height (<140 cm), inconsistent response to comparable questions in the survey (e.g., inconsistent response to questions about marital status and living area), and incorrect response to a staged question used to identify fraudulent responses (choose the third largest number from the following five numbers). Of the remaining 27,036 participants, 6,641 who indicated that their job primarily involved physical labor were excluded because these workers are unable to telecommute and were not relevant to the study’s aim to investigate the effect of telecommuting on office workers. Data from the remaining 20,395 (9,964 males and 10,431 females) participants who indicated that their work was primarily desk work or primarily involved interpersonal communication were used for analysis.

This study was approved by the ethics committee of XXXX. Informed consent was obtained via a form on the survey website.

### Assessment of telecommuting frequency and preference

The participants were asked to answer the following question about their telecommuting status: “Do you currently telecommute?” by selecting from “More than 4 days per week,” “More than 2 days per week,” “Less than 1 day per week,” and “Hardly ever.”

The participants indicated their preference for telecommuting by selecting from “I want to telecommute as much as possible,” “I prefer to telecommute,” “Either is fine with me,” “I’d rather work in the company office,” and “I’d prefer to work in the company office as much as possible.”

### Assessment of psychological distress

We examined psychological distress using Kessler 6 (K6) ^16^ The Japanese version of the K6 was validated in a previous study.^19^ The K6 was initially devised to identify patients with mental disorders, such as depression and anxiety, and is additionally used to determine the degree of mental problems, including psychological stress. The K6 contains six questions, each with scores ranging from 0 (never) to 4 (always), depending on the frequency the event described in each question was experienced in the past 30 days. A higher total score indicates a greater potential for depression or anxiety disorder, with a score ≥5 suggesting a possible depression or anxiety problem and a score ≥13 suggesting suspected depression or anxiety disorder. We used a K6 score ≥5 to indicate the presence of mild psychological distress.

### Other covariates

We used socioeconomic, lifestyle, health-related, and occupation-related factors as confounding factors. Socioeconomic factors were age, sex, marital status (married, unmarried, bereaved/divorced), occupation (primarily desk work, jobs primarily involving interpersonal communication), education level, and equivalent income (household income divided by the square root of household size). Lifestyle factors were smoking status (never; quit smoking more than one year ago; quit smoking within the past year; started smoking less than one year ago; smoking for more than one year), alcohol consumption (6–7 days a week; 4–5 days a week; 2–3 days a week; less than 1 day a week; hardly ever), exercise habit (number of days per week during which participant exercised for ≥30 min; free response), time spent on one-way commute and overtime work hours per day. Health-related factors were self-rated health and the presence of any disease necessitating repeated hospital visits or treatment (“I do not have such a disease;” “I am continuing with hospital visits and treatment as scheduled;” “I am unable to continue with hospital visits and treatment as scheduled”).

Further we also used the cumulative COVID-19 infection rate one month before initiation of the survey in the participants’ prefecture of residence as a community-level variable.

### Statistical analysis

We estimated the age-sex-adjusted and multivariate-adjusted odds ratios (ORs) of psychological distress associated with telecommuting frequency. The analyses were stratified by telecommuting preference after confirming the presence of a significant interaction between telecommuting frequency and preference in a preliminary analysis. We accounted for differences in residential area by adopting a multilevel logistic model nested in the prefecture of residence. The multivariate model included sex, age, marital status, job type, equivalent household income, education level, self-rated health, the presence of any disease necessitating repeated hospital visits or treatment, smoking status, alcohol consumption, exercise habit, time spent on one-way commute and overtime work hours per day. The COVID-19 infection rate by prefecture was used as a prefecture-level variable in all comparisons. *p*<0.05 was used to indicate statistical significance. All analyses were performed in Stata (Stata Statistical Software: Release 16; StataCorp LLC, TX, USA).

## Results

Table 1 summarizes the participants’ baseline characteristics. Among the 20,395 respondents, 2,537 (12%) telecommuted more than 4 days per week, 1,391 (7%) telecommuted more than 2 days per week, 1,343 (7%) telecommuted less than 1 day per week, and 15,124 (74%) hardly ever telecommuted. Regarding telecommuting preference, 7,264 (35.2%) participants indicated that they preferred to work in the office and 7,390 (36.2%) stated they preferred to telecommute. Thirty-nine percent of participants were identified as having mild psychological distress.

**Table 1.** Participants’ basic characteristics

|  | Telecommuting frequency |  |  |  |
| --- | --- | --- | --- | --- |
|  | ≥4 days per week<br>n=2537 | 2-3 days per week<br>n=1391 | ≤1 day per week<br>n=1343 | Hardly ever<br>n=15124 |
| Age, years, mean (SD) | 49.5 (10.0) | 48.8 (10.3) | 47.8 (10.4) | 46.4 (10.6) |
| Sex, male | 1417 (55.9%) | 806 (57.9%) | 831 (61.9%) | 6910 (45.7%) |
| Marital status, married | 1287 (50.7%) | 868 (62.4%) | 876 (65.2%) | 8540 (56.5%) |
| Education |  |  |  |  |
| Junior high school | 22 (0.9%) | 10 (0.7%) | 4 (0.3%) | 137 (0.9%) |
| High school | 429 (16.9%) | 163 (11.7%) | 199 (14.8%) | 3637 (24.0%) |
| Vocational school/college, university, graduate school | 2086 (82.2%) | 1218 (87.6%) | 1140 (84.9%) | 11350 (75.0%) |
| Annual equivalent household income (JPY) |  |  |  |  |
| 500000-2650000 | 895 (35.3%) | 286 (20.6%) | 253 (18.8%) | 4697 (31.1%) |
| 2650000-4500000 | 651 (25.7%) | 404 (29.0%) | 366 (27.3%) | 5026 (33.2%) |
| >4500000 | 991 (39.1%) | 701 (50.4%) | 724 (53.9%) | 5401 (35.7%) |
| Job type |  |  |  |  |
| Primarily desk work | 2042 (80.5%) | 1058 (76.1%) | 952 (70.9%) | 9416 (62.3%) |
| Jobs primarily involving interpersonal communication | 495 (19.5%) | 333 (23.9%) | 391 (29.1%) | 5708 (37.7%) |
| Self-rated health, very good/good | 1372 (54.1%) | 778 (55.9%) | 731 (54.4%) | 7525 (49.8%) |
| Presence of any disease necessitating regular hospital visits or treatment | 965 (38.1%) | 531 (38.2%) | 494 (36.8%) | 5239 (34.7%) |
| Current smoker | 634 (25.0%) | 379 (27.2%) | 392 (29.2%) | 3677 (24.3%) |
| Alcohol consumption, 6–7 days a week | 598 (23.6%) | 339 (24.4%) | 309 (23.0%) | 3085 (20.4%) |
| Exercise habit, more than 1 days a week | 709 (27.9%) | 417 (30.0%) | 414 (30.8%) | 3102 (20.5%) |
| Time spent on one-way commute (excluding time spent at home), more than 2 hours | 49 (1.9%) | 63 (4.5%) | 48 (3.6%) | 307 (2.0%) |
| Overtime work, more than 2 hours | 219 (8.6%) | 162 (11.6%) | 157 (11.7%) | 1394 (9.2%) |
| Telecommuting preference |  |  |  |  |
| Prefer not to telecommute | 133 (5.2%) | 186 (13.4%) | 332 (24.7%) | 6613 (43.7%) |
| No preference for or against telecommutin | 372 (14.7%) | 331 (23.8%) | 419 (31.2%) | 4619 (30.5%) |
| Prefer to telecommute | 2032 (80.1%) | 874 (62.8%) | 592 (44.1%) | 3892 (25.7%) |
| Psychological distress (K6 ≥5) | 976 (38.5%) | 533 (38.3%) | 522 (38.9%) | 5977 (39.5%) |

Table 2 displays the odds ratios (ORs) of mild psychological distress associated with telecommuting frequency stratified by telecommuting preference. Among those who preferred not to telecommute, the OR of psychological distress increased as the telecommuting frequency increased. Compared to those who rarely telecommuted, the multivariate OR of psychological distress among those who telecommuted four or more days per week was 1.87 (95% CI: 1.29-2.73, p=0.001). Conversely, among those who preferred to telecommute, the OR of psychological distress decreased as the telecommuting frequency increased. Compared to those who rarely telecommuted, the multivariate OR of psychological distress among those who telecommuted four or more days per week was 0.67 (95%CI: 0.58-0.78, p<0.001). Meanwhile there was no association between telecommuting frequency and psychological distress among those with no preference for or against telecommuting.

**Table 2.** Odds ratio of psychological distress associated with telecommuting frequency stratified by telecommuting preference

|  | Age-sex adjusted |  |  |  | Multivariate<sup>a</sup> |  |  |  |
| --- | --- | --- | --- | --- | --- | --- | --- | --- |
|  | OR | 95%CI |  | p | OR | 95%CI |  | p |
| Among subjects who preferred not to telecommute |  |  |  |  |  |  |  |  |
| Telecommuting frequency |  |  |  |  |  |  |  |  |
| More than 4 days per week | 1.74 | 1.22 | 2.48 | 0.002 | 1.87 | 1.29 | 2.70 | 0.001 |
| More than 2 days per week | 1.40 | 1.03 | 1.91 | 0.033 | 1.36 | 0.98 | 1.87 | 0.064 |
| Less than 1 day per week | 1.27 | 1.00 | 1.61 | 0.046 | 1.27 | 0.99 | 1.62 | 0.060 |
| Hardly ever | reference |  |  |  | reference |  |  |  |
| Among subjects who had no preference for or against telecommuting |  |  |  |  |  |  |  |  |
| Telecommuting frequency |  |  |  |  |  |  |  |  |
| More than 4 days per week | 1.10 | 0.87 | 1.37 | 0.427 | 1.15 | 0.89 | 1.47 | 0.286 |
| More than 2 days per week | 0.95 | 0.74 | 1.21 | 0.657 | 0.93 | 0.72 | 1.20 | 0.573 |
| Less than 1 day per week | 0.95 | 0.76 | 1.18 | 0.627 | 1.03 | 0.83 | 1.29 | 0.773 |
| Hardly ever | reference |  |  |  | reference |  |  |  |
| Among subjects who preferred to telecommute |  |  |  |  |  |  |  |  |
| Telecommuting frequency |  |  |  |  |  |  |  |  |
| More than 4 days per week | 0.65 | 0.58 | 0.72 | <0.001 | 0.67 | 0.58 | 0.78 | <0.001 |
| More than 2 days per week | 0.68 | 0.58 | 0.79 | <0.001 | 0.73 | 0.62 | 0.86 | <0.001 |
| Less than 1 day per week | 0.75 | 0.63 | 0.89 | 0.001 | 0.79 | 0.66 | 0.95 | 0.014 |
| Hardly ever | reference |  |  |  | reference |  |  |  |

<sup>a</sup> The multivariate model included sex, age, marital status, job type, equivalent household income, education level, self-rated health, the presence of any disease necessitating regular hospital visits or treatment, smoking status, alcohol consumption, exercise habit, time spent on one-way commute, overtime work hours per day, and the COVID-19 infection rate by prefecture as a prefecture-level variable.

## Discussion

We found that the relationship between telecommuting frequency and psychological distress during the COVID-19 pandemic differed depending on telecommuting preference. Workers who preferred to telecommute experienced less psychological distress with increasing telecommuting frequency, while those who preferred not to telecommute experienced more psychological distress with increasing telecommuting frequency.

The present study revealed that there is a mismatch between telecommuting preference and actual telecommuting. The ongoing COVID-19 pandemic has forced many workers to adopt telecommuting involuntarily due to government directives. Our findings provide insight into the health impact of telecommuting in terms of workers’ preference.

We found that the direction of the association between telecommuting frequency and psychological distress was completely different depending on telecommuting preference. This finding is in line with previous studies that have noted that telecommuting has both positive and negative effects on mental health. Tavares et al. argued that there are two positive effects of telecommuting. First, the flexibility provided by telecommuting gives workers enhanced control over their lives, decreases family conflicts, and improves work-life balance, thereby reducing workers’ exposure to a number of stressors. Second, telecommuting provides workers with resources to deal with stressors, thereby preventing the adverse effects of stress on health. However, some negative effects of the increased flexibility provided by telecommuting include stress due to family responsibilities, inability to separate work and home life, and possible family conflict. Further, telecommuting has greater potential to bring about overwork, tight deadlines, intense and long working hours, little rest time, and the inability to switch off, all of which are associated with poor mental health, fatigue, and poor health.^6^ Our findings suggest that the negative effects may be stronger among workers who prefer not to telecommute.

There are several possible reasons why workers may prefer not to telecommute, although there is current little evidence on telecommuting preference and choice. First, workers may prefer not to telecommute if they do not have a suitable environment or setup for work at home. Appropriate work environments such as a desk designed for office work, a quiet room, and comfortable temperature, humidity, and light intensity are basic requirements for office workers. These affect not only a worker’s performance, but also their health, with poor conditions being a potential initiating or exacerbating factor for back pain and eye strain.^10–12^

Second, in addition to the physical environment, workers’ cohabitation status may affect their telecommuting preference. Workers may have small child who require constant care, or live with an elderly person who requires nursing care. Providing such care would typically take up a substantial amount of time, and going to work may offer these workers some respite that would no longer be available if they were to telecommute. In addition, attendance at nursery schools and elderly care facilities was likely interrupted during the COVID-19 pandemic, ^20–25^ making such care impossible to avoid while telecommuting.

Third, telecommuting reduces communication and, for this reason, is considered undesirable for jobs that place importance in communication with colleagues and customers. In these circumstances, being forced to telecommute may increase mental strain related to stress and job control.^26,27^ Tentative evidence suggests that flexible work interventions that give workers more control and choice are likely to have a positive effect on health outcomes.^28^ Telecommuting during the COVID-19 pandemic is thought to have greatly impaired workers’ job control because it was introduced without their agreement.

These findings have several implications for public health. First, for those who prefer to work from home, telecommuting has the potential to reduce psychological distress. Thus, company strategies that make telecommuting accessible, the establishment of telecommuting environments, and support to help workers feel they have control over their jobs can facilitate this positive impact. Second, those who prefer not to telecommute require support to reduce their mental burden. As telecommuting is increasingly being introduced as a measure to deter infection, it is necessary to give workers the option to work in the company office wherever possible and to monitor and support their mental burden when working from home. It would also be useful to identify the reasons why workers prefer not to telecommute and to provide support to reduce any burden where possible.

This study has several limitations. First, since this study is a survey of Internet monitors, selection bias was unavoidable. However, to reduce bias as much as possible, we selected subjects according to region, occupation, and prefecture based on infection rates. Second, we did not obtain any detailed information about participants’ industry or job description. Telecommuting frequency and preference may be related to whether the work is performed at a site that makes telecommuting impossible, such as a factory, or at an office with sufficient Internet connection. Third, telecommuting preference may be related to workers’ health. Workers who are in poor health, including those with mental illness, may be more likely to choose to work from home.

In conclusion, this study showed that the association between psychological distress and telecommuting frequency differed depending on telecommuting preference during the COVID-19 pandemic. Workers who preferred to telecommute experienced less psychological distress with increasing telecommuting frequency, while those who preferred not to telecommute experienced more psychological distress with increasing telecommuting frequency. It is thus important to consider workers’ telecommuting preference when deciding their telecommuting frequency to limit psychological distress.

## Data Availability

Data not available due to ethical restrictions

## Acknowledgements

The current members of the CORoNaWork Project, in alphabetical order, are as follows: Dr. Yoshihisa Fujino (present chairperson of the study group), Dr. Akira Ogami, Dr. Arisa Harada, Dr. Ayako Hino, Dr. Hajime Ando, Dr. Hisashi Eguchi, Dr. Kazunori Ikegami, Dr. Kei Tokutsu, Dr. Keiji Muramatsu, Dr. Koji Mori, Dr. Kosuke Mafune, Dr. Kyoko Kitagawa, Dr. Masako Nagata, Dr. Mayumi Tsuji, Ms. Ning Liu, Dr. Rie Tanaka, Dr. Ryutaro Matsugaki, Dr. Seiichiro Tateishi, Dr. Shinya Matsuda, Dr. Tomohiro Ishimaru, and Dr. Tomohisa Nagata. All members are affiliated with the University of Occupational and Environmental Health, Japan.

